# The importance of supplementary immunisation activities to prevent measles outbreaks during the COVID-19 pandemic in Kenya

**DOI:** 10.1101/2020.08.25.20181198

**Authors:** CN Mburu, J Ojal, R Chebet, D Akech, B Karia, J Tuju, A Sigilai, K Abbas, M Jit, S Funk, G Smits, PGM van Gageldonk, FRM van der Klis, C Tabu, DJ Nokes, LSHTM CMMID COVID-19 Working Group, JAG Scott, S Flasche, IMO Adetifa

**Author notes:** these authors contributed equally, and their order was chosen at random (S Flasche, IMO Adetifa). Corresponding author: Caroline Mburu, KEMRI-Wellcome Trust Research Programme, Kilifi, Kenya., Alternate corresponding author; Ifedayo Adetifa KEMRI-Wellcome Trust Research Programme, Kilifi, Kenya.

## Abstract

**Background:** The COVID-19 pandemic has disrupted routine measles immunisation and supplementary immunisation activities (SIAs) in most countries including Kenya. We assessed the risk of measles outbreaks during the pandemic in Kenya as a case study for the African Region.

**Methods:** Combining measles serological data, local contact patterns, and vaccination coverage into a cohort model, we predicted the age-adjusted population immunity in Kenya and estimated the probability of outbreaks when contact-reducing COVID-19 interventions are lifted. We considered various scenarios for reduced measles vaccination coverage from April 2020.

**Findings:** In February 2020, when a scheduled SIA was postponed, population immunity was close to the herd immunity threshold and the probability of a large outbreak was 22% (0-46). As the COVID-19 restrictions to physical contact are lifted, from December 2020, the probability of a large measles outbreak increased to 31% (8-51), 35% (16-52) and 43% (31-56) assuming a 15%, 50% and 100% reduction in measles vaccination coverage. By December 2021, this risk increases further to 37% (17-54), 44% (29-57) and 57% (48-65) for the same coverage scenarios respectively. However, the increased risk of a measles outbreak following the lifting of restrictions on contact can be overcome by conducting an SIA with ≥ 95% coverage in under-fives.

**Interpretation:** While contact restrictions sufficient for SAR-CoV-2 control temporarily reduce measles transmissibility and the risk of an outbreak from a measles immunity gap, this risk rises rapidly once physical distancing is relaxed. Implementing delayed SIAs will be critical for prevention of measles outbreaks once contact restrictions are fully lifted in Kenya.

**Funding:** The United Kingdom’s Medical Research Council and the Department for International Development

## Background

The SARS-CoV-2 pandemic has interrupted large parts of social interaction, the economy and important health services in Kenya and around the world.^1,2^ As of mid-July 2020, the incidence of COVID-19 cases continues to rise across most of Africa, implying that the current mitigation measures need to be maintained or extended for a period at least until the pandemic peaks.^3^

Despite the World Health Organization (WHO) advisory to sustain routine immunization (RI), vaccine coverage has temporarily declined in many countries including Kenya that reports a 33% disruption of RI.^4-7^ Following guidance from the WHO, all countries suspended planned supplementary immunisation activities (SIAs) for measles.^6-8^ Measles control in Kenya is achieved by a delivery of a first dose of Measles Containing Vaccine (MCV1) at 9 months, and a second dose (MCV2) from 18 months. SIAs, first introduced in 2002 are conducted periodically among children <5 years or <15 years for accelerated control of measles.^9^ Based on accumulation of susceptible children, timing of such campaigns has typically been chosen to close immunity gaps in time to prevent potentially large measles outbreaks. Measles SIA originally due in 2019 was rescheduled for February 2020 due to a shortfall in funding and postponed again following the COVID-19 pandemic.

Following identification of the first COVID-19 case on March 13, 2020, Kenya imposed various mitigation measures: ban on large gatherings, suspension of international flights, closure of bars, cessation of movement from hotspot counties, restriction of restaurant operating hours and a nationwide curfew from 7pm to 5 am. While it is plausible that these physical distancing and lock down measures may reduce the risk of measles outbreaks, they are temporary and may be associated with rebound risk periods.

The availability of recent measles serological data provided the opportunity to use Kenya as a case study to estimate the impact of reduced measles vaccination coverage and suspended SIAs due to COVID-19 on the risk of measles outbreaks.

## Methods

This study used a cohort mathematical model that combined measles serological data from a survey in October 2019, local contact patterns, and vaccination coverage estimates.

### Serological data

We estimated measles immunity profile in children using samples collected during serosurveys of the Pneumococcal Conjugate Vaccine Impact Study (PCVIS).^10^ The participants were a randomly selected sample of children in ten age strata (0, 1, 2, 3, 4, 5, 6, 7, 8-9 and 10-14 years) resident in Kilifi Health and Demographic Surveillance System (KHDSS) in Kilifi, Kenya.^11^ In the 2019 serosurvey, 468 participants were recruited of which 214 randomly selected blood samples had been tested for presence of measles antibodies before COVID-19 disruptions to work. The samples were collected in July (107), August (73), September (33) and October (1) 2019. Measles immunoglobulin G (IgG) antibodies were detected using a fluorescent-bead-based multiplex immunoassay and antibody concentrations ≥0.12 IU/ml were considered protective against measles.^12^

For projections, we assumed these results reflected measles immunity in Kilifi in August 2019, and assumed 96% of persons >15 years had protective measles antibodies concentrations, similar to findings in adults in Nairobi in 2007-2009^13^ (Table 1). We also assumed no protection from maternal immunity since none of the infants <9 months old had protective antibodies.

### Vaccination coverage

MCV1 national coverage in Kenya has been between 75% and 80% since it was introduced in 1985.^14^ MCV2 was introduced into the RI programme in Kenya in 2013 and coverage rose to 45% in 2018 after introduction.^9^ The last measles SIA in children aged 9 months to 14 years took place in 2016 and achieved 95% coverage.^15^

We assumed national MCV1 and MCV2 coverage were 79% and 45% respectively in 2018, and that these stayed at the same level from August 2019 until the end of March 2020 when COVID-19 contact restrictions were introduced in Kenya. From April 2020, we explored the following routine vaccination coverage scenarios alongside a suspended SIA

A. routine vaccination coverage remained the same, or
B. routine vaccination coverage reduced by 15% for both MCV1 and MCV2, or
C. routine vaccination coverage reduced by 50% for both MCV1 and MCV2, or
D. routine vaccination was suspended

### Contact matrix

We used an age-mixing matrix from a study conducted in Kilifi, Kenya.^16^ The matrix which consisted of the number of contacts between six different age groups was used in generating contact-adjusted immunity estimates.

### Projecting immunity

We adapted a static cohort model of measles immunity^17^ to estimate age stratified population immunity profile in Kilifi by combining recent measles serological data with new vaccine-derived immunity during the prediction period using the local vaccination schedule, MCV1 and MCV2 uptake, and vaccine efficacy. We assumed waning immunity or additional acquired immunity from natural exposure, and demographic changes in the short time frame considered were negligible. Hence, the key mechanisms of the projection model were that individuals are born at a constant rate, gain immunity through vaccination at the recommended age and at the observed coverage, and grow older.

In extrapolating immunity for young infants under 9 months old, maternal immunity was assumed to be the same as the observed data. For ages 9 months to 17 months, immunity was estimated in accordance with the assumed MCV1 vaccination uptake and a vaccine effectiveness of 93%. For those ≥18 months, we estimated the immunity based on the assumed uptake of MCV2 and the same vaccine effectiveness. We aggregated projected immunity to age groups given by contact data and weighted each age group according to population estimates before averaging them to estimate overall immunity. We did not explicitly model MCV2 delivery but rather assumed that the MCV1 effectiveness is an average of MCV1 and MCV2 efficacy weighted by proportion of children who receive MCV1 only or both doses. The underlying assumption here was that the same children who received MCV2 had also received MCV1. We predicted age stratified and population level immunity until December 2021.

To derive a contact-adjusted estimate for the proportion of the population who are immune to measles, the predicted age stratified immunity profile was weighted by age stratified social contact patterns observed in Kilifi. This method has been previously shown to yield robust projections for measles immunity to transmission in the population.^17^

The herd immunity threshold for measles during the COVID-19 pandemic was calculated assuming an R_0_ of 12 to 18 with a median of 14^18^ and that COVID-19 prescribed contact restrictions caused a 50% reduction in measles transmissibility similar to the observed reduction in physical contacts in Kenya.^19^ The herd immunity threshold is calculated as 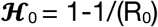

### Quantitative impact of outbreak risk

We obtained a crude estimate of the outbreak risk using the predicted immunity and herd-immunity threshold. The probability of a large outbreak, p, sparked by a single infected individual was given by p = 1-(1/R)^I0^ where I0 is the initial number infected and R is the effective reproductive number. R<1 implies that probability, p, is negative which is defined to be 0 for no outbreak.

### The effectiveness of a post-lockdown SIA in reducing outbreak risk

We assessed the impact of SIAs in two age categories; 9 months to 5 years and 9 months to 15 years, by predicting the post-SIA immunity profile and the corresponding risk for a large measles outbreak. We simulated SIAs in either October 2020, December 2020 or December 2021, assumed a coverage of 95% similar to the most recent national SIA in 2016,^15^ and applied vaccine efficacy of MCV1. The SIA was simulated by reducing the age specific pool of susceptible by the effective coverage of the SIA.

### Uncertainty analyses

We assessed the sensitivity of our findings to uncertainty inherent in several of our assumptions via probabilistic re-sampling. We included uncertainty for population immunity profile, combined MCV1 and MCV2 vaccine effectiveness, and MCV1 and MCV2 coverage (Table 1). As part of each parameter bootstrap, we also bootstrapped participants of the serological survey and hence the age stratified population immunity at the start of the simulation. We present median estimates including uncertainty quantified as per the 95% quantiles of the bootstrap analyses.

### Sensitivity analyses

We conducted a sensitivity analysis to assess the impact of a delay in receipt of MCV1 on outbreak probability. We delayed the age of receipt of MCV1 in our model by three months as reported for delayed vaccination in Kilifi.^20^ We also predicted unadjusted population immunity in Kilifi and estimated the corresponding probability of a large outbreak

All analyses were done in R^21^ and are available on github at: https://github.com/CarolineNM/ncov_measles_Kenya

**Table 1:** Model parameters. An overview of the key model parameter assumptions and their sources. Parameter ranges are those used in the sensitivity analysesSource.

| Parameter | Value (95% quantiles) | Source |
| --- | --- | --- |
| Vaccine schedule | MCV1: 9 months<br>MCV2: 18 months | 22 |
| Vaccine effectiveness<br>(Beta distributed) | MCV1: 85% (80 - 90%)<br>+MCV2: 98% (95 - 100%)<br>Combined effectiveness<br>93% (88-96%) | 23,24 |
| Age-Immunity profile in <15y old<br>(Bootstrapped from data) | Observed in 2019 | 10 |
| Proportion immune among >15y old<br>(Beta distributed) | 96% immune (90 - 99%) | 13 |
| Vaccine coverage Aug 2019 to March 2020<br>(assumed to be same as in 2018)<br>(Beta distributed) | MCV1: 79% (75-85%)<br>MCV2: 45% (40-50%) | 9,14,20 |
| Vaccine coverage from April 2020 | MCV1 & MCV2 0%, 15%,<br>50% or 100% reduced | assumption |
| R<sub>0</sub> measles<br>(Log-normally distributed) | 14 (12 - 18) | 18 |
| Reduction in contacts during COVID-19 | 50% | 19 |
| Age demographics | from KHDSS in 2019 | 11 |
| Social mixing matrix | from 2011/12 | 16 |

## Results

### Measles seroprevalence in Kilifi in late 2019

Proportion of MCV1-eligible children with protective measles antibody concentrations was high in late 2019 (Figure 1). All 25 children over 10 years old had protective levels. Similarly, 77 of 82 (94%) 5-9-year-olds were immune. Among children under 5 years eligible for MCV1, 86 of 97 (89%) were immune but none of the 10 children who were age ineligible for vaccination had protective antibodies.

**Figure 1.**
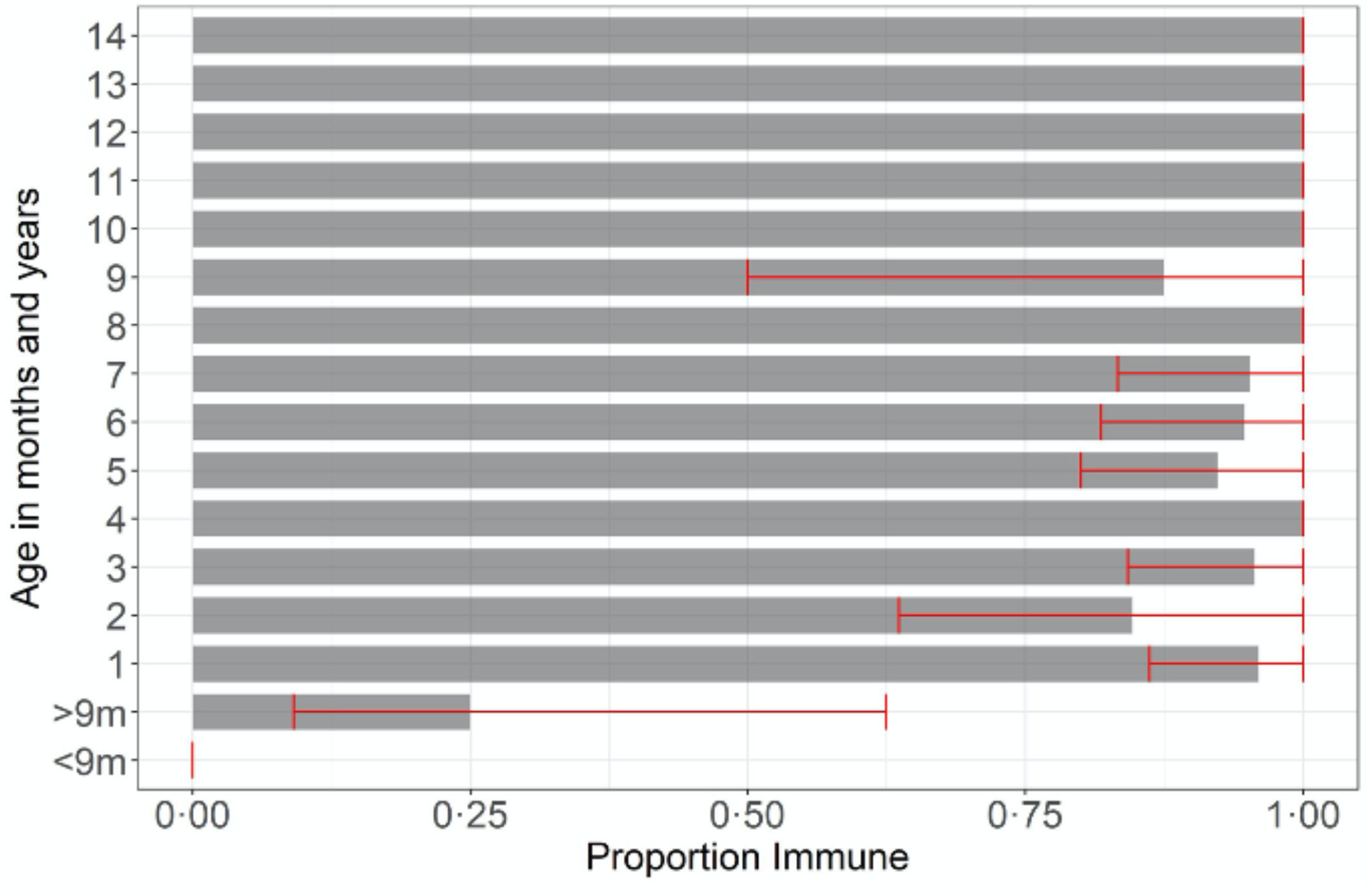
Age-stratified population immunity profile. Estimated age-stratified proportion of the Kilifi County population who were immune to measles infection in August 2019 from data. Antibody concentrations ≥0-12 IU/ml were defined as protective. Confidence bounds displayed (in red) are the 95% quantiles of a nonparametric bootstrap that is used to propagate uncertainty into the modelling framework. MCV1 is recommended to be administered at 9 months as per the Kenyan immunisation schedule and MCV2 from 18 months

### Age adjusted immunity

We estimate that in late 2019, population immunity adjusted for age-differences in social contacts was 91% (87-93). Predicted proportion immune was unchanged in February 2020, at the time of originally planned SIA.

Following the start of COVID-19 pandemic and restriction measures that caused a decrease in vaccination coverage, we estimate that population immunity decreased quickly, depending on extent of reduction in vaccination coverage. If vaccine coverage is reduced by 15% from April 2020 onwards, contact-adjusted population immunity would decline to 90% (87-92) by December 2020 and 89% (87-92) by December 2021. A 50% reduction in vaccination coverage would lead to a more rapid decline in this immunity to 89% (86-91) in December 2020 and 87% (84-89) in December 2021(Figure 2)

**Figure 2.**
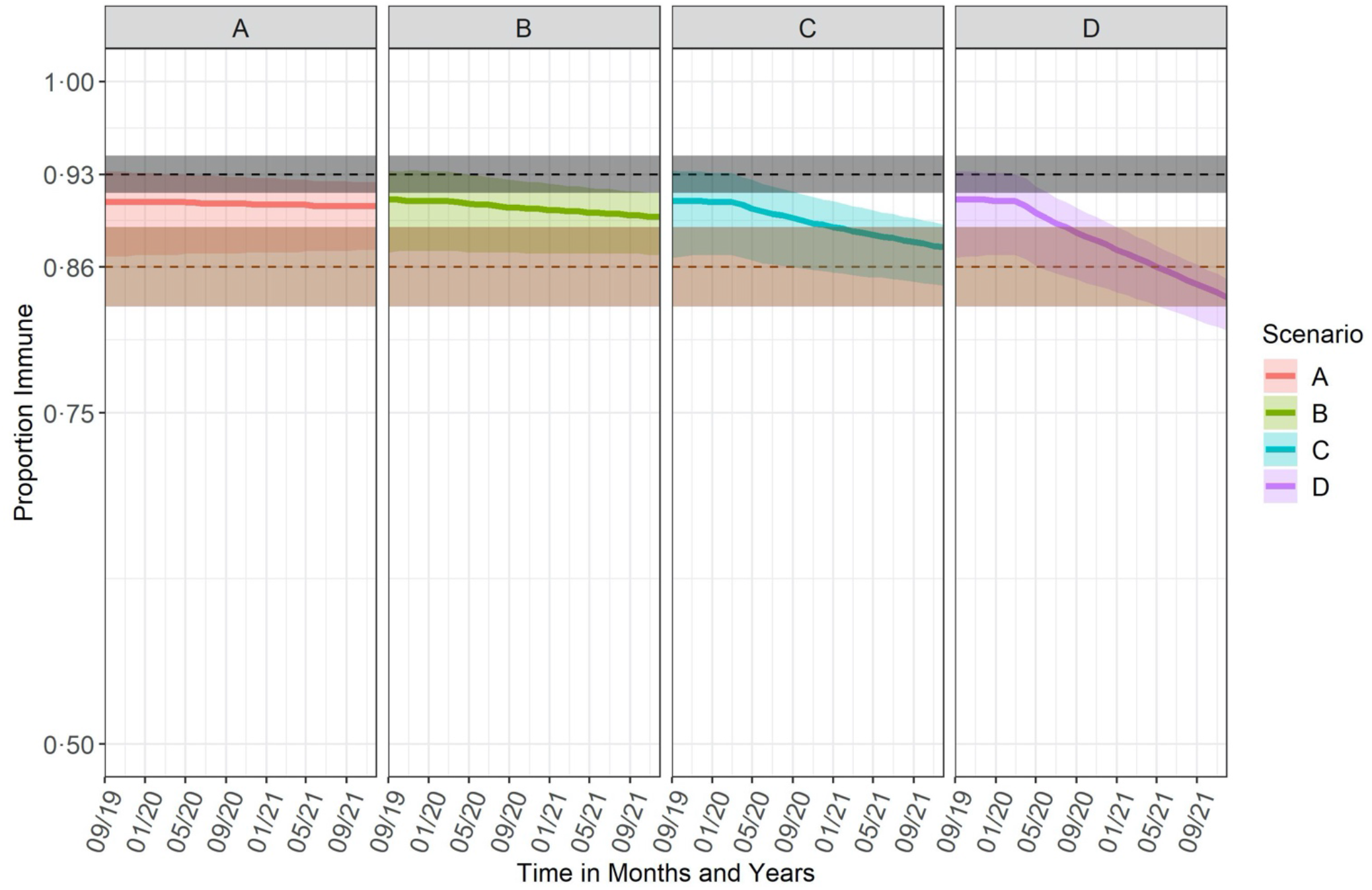
Monthly projected age adjusted immunity profiles from September 2019 to December 2021. The changes in coverage took effect in April 2020. The black dotted line shows the herd immunity threshold for measles before the COVID-19 physical distancing measures, 0·93 (0·92 to 0·94) and the brown dotted line shows the herd immunity threshold during COVID-19 physical distancing measures, 0·86[0·83-0·89], assuming the lockdown measures are still in effect. The bold lines and shaded region in each scenario i.e. A. No reduction, B. 15% reduction, C. 50% reduction and D. 100% reduction indicate the median estimates and the uncertainty of the predicted immunity quantified as the 95% quantiles of the bootstrap analysis. There was a quick decline of predicted immunity over the study period that was based on assumed reduction in routine coverage

### Age adjusted immunity vs herd immunity threshold

A basic reproduction number of 14 (12-18) implies a herd immunity threshold of 93% (82-94) and if, as a result of social distancing, measles transmission is reduced by 50%, herd immunity threshold drops to 86% (83-89) as seen in figure 2.

Before contact restrictions came into effect in April 2020, age-adjusted immunity was slightly below herd immunity threshold: in 10% of simulations this immunity was below the herd immunity threshold (Figure 3). Reduction in herd immunity threshold temporarily mitigated immediate risk for measles outbreak as in all simulations immunity in April 2020 was above the reduced transmission herd immunity threshold.

**Figure 3.**
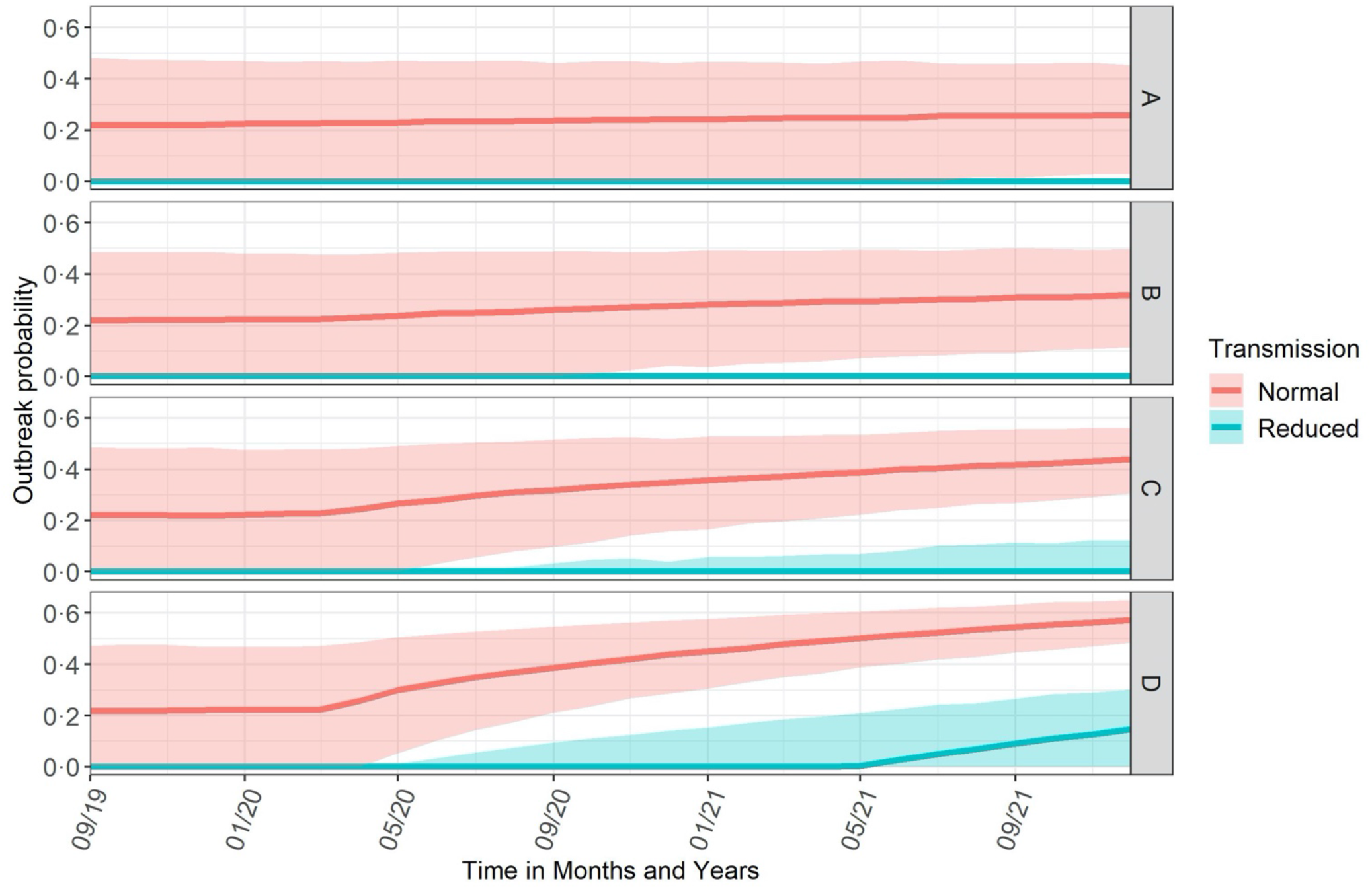
Probability of a large measles outbreak sparked by a single infected individual. Outbreak probability was calculated using the predicted immunity and herd immunity threshold before (red) and during (green) COVID-19 movement restriction measures. Zero probability indicates no possibility of an outbreak. The bold lines and shaded region in each scenario i.e. A. No reduction, B. 15% reduction, C. 50% reduction and D. 100% reduction indicate the median estimates of outbreak risk and the uncertainty quantified as the 95% quantiles of the bootstrap analysis. The risk of a large measles outbreak from the introduction of a single infectious individual increased quickly based on the level of impairment of routine vaccination coverage

Depending on vaccination coverage maintained during COVID-19 pandemic, population immunity may decline quickly in young children (<2 years). By April 2020, age-adjusted immunity fell below the normal transmission herd immunity threshold in more than 95% of simulation under scenario C and D respectively. (Figure S2).

Similarly, risk of a large measles outbreak from introduction of a single infectious individual increased quickly if routine vaccination coverage was impaired (Figure 3). If in October 2020, measles transmissibility was similar to that pre-COVID-19 and routine measles coverage since April 2020 reduced by 15%, 50% or 100%, we estimate probability for a large measles outbreak as 26% (8-49), 33% (11-52), 40% (24-56) respectively in the age-adjusted analysis. By the end of the year, the risk would increase to 28% (4-49), 35% (16-52) and 44% (29-57) respectively.

### Effectiveness of a post-lockdown SIA

SIAs in 9 months to 5-year-old children or 9 months to 15-year-olds immediately after lifting transmission-reducing COVID-19 mitigation measures can substantially reduce outbreak risk (Figure 4).

**Figure 4.**
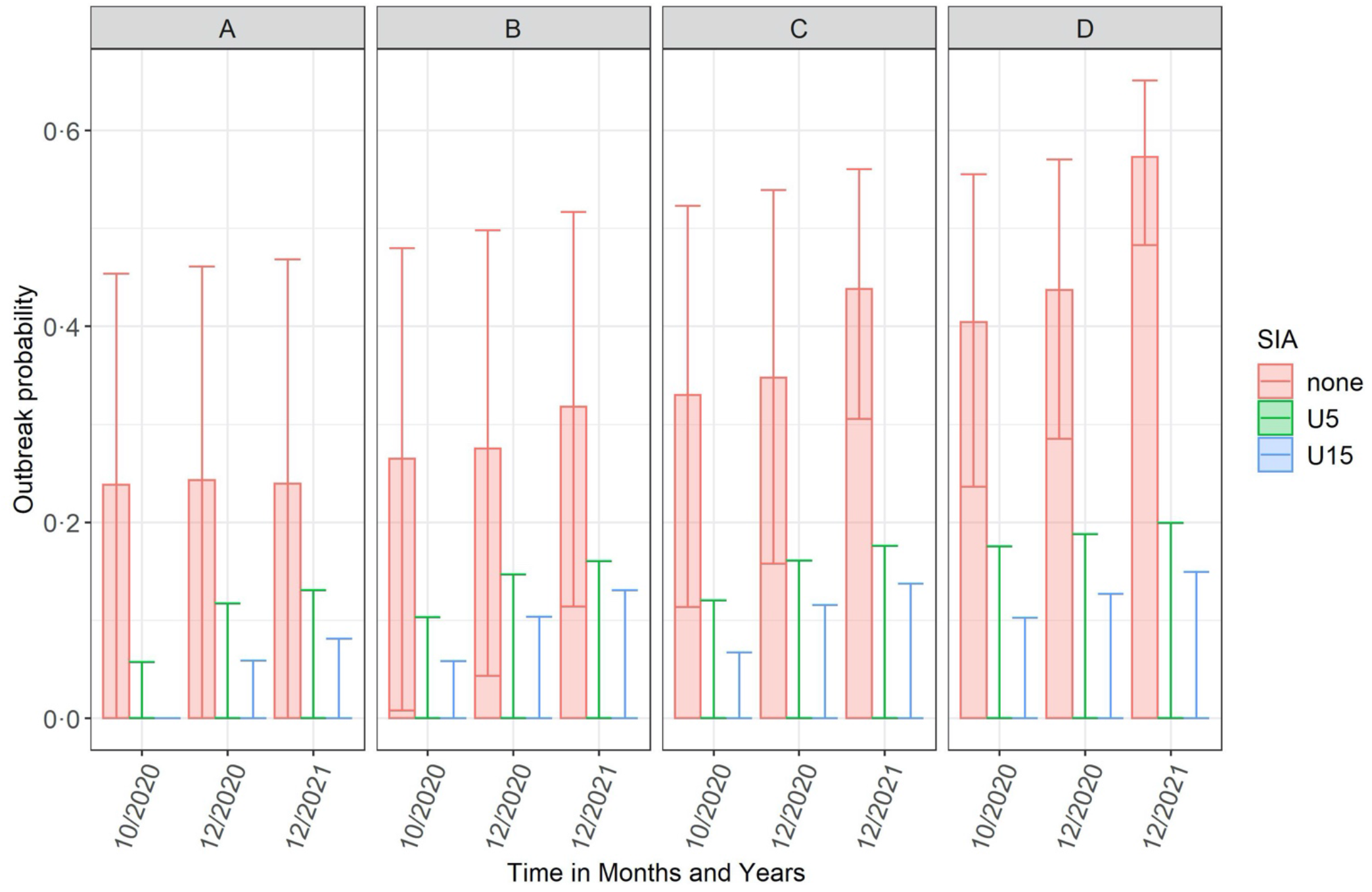
Probability of a single infectious person seeding a large outbreak before (none) and after implementing an SIA in children 9 months to 5 years old (U5) and in 9 months to 15 years old (U15) at different timepoints. Outbreak probability was calculated by comparing the proportion immune with the herd immunity threshold. The shaded area is the median estimate of the outbreak risk and the error bars indicate the uncertainty in outbreak risk quantified as the 95% quantiles of the bootstrap analysis. In all the scenarios, i.e. A. No reduction, B. 15% reduction, C. 50% reduction and D. 100% reduction, the risk of a large measles outbreak would be largely mitigated through delivery of a SIA among children <5 years old or <15 years old.

If routine measles vaccine coverage was reduced by 15%, 50% or 100% since April 2020, an SIA delivered to children 9 months to 5 years old in October 2020 with 95% coverage would reduce measles outbreak probability risk to 0% (0-11), 0% (0-12) and 0% (0-17) for coverage scenarios respectively in age-adjusted analysis. Even if RI coverage is impaired through to December 2021, the risk for a large measles outbreak would be mitigated through an SIA for under-fives if delivered before or immediately after contact restrictions are lifted (Figure 4).

### Impact of delayed vaccination on outbreak probability

Receipt of MCV delayed by 3 months in children resulted in a marginal increase in the risk of a large measles outbreak from introduction of a single infectious individual (Figure S1).

### Crude Population immunity

Predicted crude population immunity was slightly higher compared to age-adjusted immunity but followed the same declining trend over time (Figure S3). Before contact restrictions came into place, 50% of simulations were below herd immunity threshold and by June 2021 and November 2020, this immunity fell below herd immunity threshold in more than 95% of simulations under scenario C and D respectively (Figure S2).

## Discussion

Our analysis suggests a decline in population immunity during COVID-19 pandemic will result in an increased risk of a measles outbreak depending on the extent to which routine vaccination coverage is reduced. We estimated the probability of a large measles outbreak from the introduction of a single infectious individual to be 26% (8-49), 33% (11-52), 40% (24-56) in October 2020 assuming a 15%, 50% or 100% reduction in routine measles vaccination coverage respectively since April 2020. This risk, which will increase to 28% (4-49), 35% (16-52) and 44% (29-57) by the end of the year will be greatly reduced if an SIA among children <5 years old is conducted before or immediately after all COVID-19 related restrictions on physical contact are lifted.

We based our analysis on an immunity model that combined serological data and age-specific mixing patterns in Kenya. Combining the two is a better strategy for predicting outbreaks as opposed to using immunity profiles alone as it allows adjustment of overall immunity by taking into account contribution of each age-group to transmission.^17^

As there is considerable uncertainty in actual reduction of routine vaccination uptake, we predicted population immunity for scenarios of routine vaccination coverage since April 2020 i.e. 15%, 50% and 100% reductions, and the corresponding outbreak risk. Our assumption of 15% reduction in vaccine coverage rates is based on reduction in vaccine clinic visits in Kilifi County (DHIS2 Routine Report) while the 50% reduction lies in the range of reported disruption in vaccination services from WHO immunisation pulse poll.^6^ We assumed a 50% reduction in measles transmissibility given that COVID-19 mitigation measures implemented on 25th March 2020 were reported to have reduced social contacts and disease transmission by the same margin.^19^ Although some restriction measures remain in place e.g. nationwide curfew, others like the partial lockdown have since been eased and ban on international flights was lifted on 1st August 2020. While the assumption of a 50% reduction in measles transmission was applicable at the beginning of the epidemic due to stringent measures imposed, current herd immunity threshold may be much higher than originally assumed but still lower than pre-COVID-19 threshold.

SIAs in Kenya are generally conducted every 2-4 years and provide a second opportunity for vaccination in children regardless of their vaccination history and are ideally timed to close immunity gaps arising from accumulation of susceptible and vaccine failures.^25^ They have been shown to be effective in increasing immunisation equity by reaching children from poor households.^26^ In February 2020, at the time of the planned national SIA, we estimated that 91 % (87-93) of the population were immune after adjusting for age-differences in social contact. This immunity which was equivalent to a 22% (0-46) probability of a large outbreak suggests the SIA would have been timely in closing immunity gaps. The risk of an outbreak which was accelerated by immunity gaps arising in children missed their routinely delivered MCV continued to increase in subsequent months following the start of COVID-19 and by December 2020, the estimated risk had increased to 28%(4-49), 35%(16-52) and 44%(29-57) assuming a 15%, 50% and 100% reduction in measles vaccination coverage respectively. Based on limited information on additional reductions in vaccination coverage as the pandemic progressed in Kenya’s devolved counties and marked reduction in vaccination services in Kenya in May 2020 compared to January and February 2020 reported in the second WHO immunisation poll, it is highly probable most areas will experience an outbreak risk of 35%(16-52) corresponding to a 50% reduction in routine coverage.

During the period of restricted movement, outbreak risks would only be experienced in the suspended RI scenario in 2021. Reduction in severity and timing of these outbreaks would be largely reduced if a measles vaccine campaign is delivered but it will also depend on time delay of catch-up campaigns and speed at which a campaign can be organised. In August 2020 for instance, an SIA would reduce outbreak risk to zero in all scenarios while in December 2020, outbreak risk would reduce to zero with an upper bound risk of 20% after delivery of SIA.

The current disruption to vaccination services will cause further delays to vaccination, which is a challenge even in normal circumstances. We had previously reported consistently poor timeliness of MCV1 vaccination across 6 different birth-cohorts (2011-2016) in Kenya.^20^ Here, a delay in age of MCV1 by 3 months resulted in a marginal increase in outbreak risk. For instance, assuming a 50% reduction in routine vaccination, a delay in vaccination would see the risk increase from 34% (15-51) to 41 % (26-57) by the end of the year. This reiterates the importance of timeliness in administration of vaccines in children as even a slight delay may cause considerable immunity gaps.

Our results emphasize the importance of maintaining high RI coverage during this pandemic because the benefits of sustaining RI services far outweighs the risks of any excess COVID-19 deaths that may arise from vaccination clinic visits.^27^ Due to the highly infectious nature of measles, massive outbreaks following disruptions to health care systems and reduced MCV1 coverage are typical. Following an Ebola outbreak in 2014-2015, Liberia and Guinea reported more than a 25% decline in MCV1 coverage.^28,29^ Reported cases also occurred in a lower age group compared to pre-Ebola period suggesting accumulation of susceptible children who missed their vaccine doses was a key contributor. Immunity gaps continued to be felt in these countries two years later even after successful implementation of SIAs.

Recently, measles outbreaks have been reported in five counties in Kenya^30^ even with COVID-19 restrictions which suggests an adverse synergistic interaction between pre-existing gaps of susceptibility due to lower vaccination coverage rates (compared to national estimates) and a precipitous drop in RI coverage during this period. These outbreaks and our results are well aligned with recent Kenya measles outbreak risk assessment report and WHO guidance on catch up vaccination and closing the immunity gaps caused by COVID-19.

As expected, majority of vaccine eligible children had protective antibody concentrations against measles but none of the 10 infants < 9 months old had protective concentrations. This suggests an extended period of susceptibility in young infants as a result of rapid maternal antibody decay. This phenomenon has been previously reported in areas where maternal immunity is increasingly from immunisation rather than natural infection.^31^

A key strength of our study is availability of recent serological data which provides an excellent means of directly estimating levels of population protection against infection and can also be used to guide post-COVID-19 SIAs. In addition, availability of an age-mixing matrix from the same area allowed us to estimate overall immunity by taking into account the level of contact between different age-groups.

Our study has a few limitations. Population immunity was only available for children <15 years but we varied observed immunity estimates in adults from a previous study in our model which resulted in a slight shift in overall immunity. The serological data, estimates and a mixing matrix used in our study may not be fully representative of the country although we utilised national estimates of vaccination coverage, which was the main driver of predicted immunity. We did not explicitly model MCV2 delivery but assumed the overall effectiveness was an average of MCV1 and MCV2 efficacy weighted by proportion of children who either receive MCV1 only or both doses. Finally, there is some uncertainty around the actual reduction in transmission due to variability in compliance with physical distancing measures in place. However, we accounted for uncertainty by varying the R0.

### Conclusions

Measles SIA originally scheduled for February 2020 in Kenya would have been well-timed as population immunity was below herd immunity threshold. Interruptions to RI since the start of COVID-19 pandemic restrictions in Kenya have now widened the measles immunity gap, but associated risk of large measles outbreaks are partially mitigated if COVID-19 contact restrictions remain in place. As these measures are almost fully lifted, we estimate that measles outbreak risks will dramatically increase, necessitating an immediate SIA’s to close the immunity gap.

## Data Availability

All analyses were done in R and the code is available on github at https://github.com/CarolineNM/ncov_measles_Kenya
The replication data and analysis scripts for this manuscript shall be made available at the KWTRP Harvard Dataverse: (https://dataverse.harvard.edu/dataverse/kwtrp). The measles serology data is part of an ongoing study and is stored under restricted access. Requests for access to the restricted dataset should be made to the Data Governance Committee of the KEMRI-Wellcome Trust Research Programme.

## Acknowledgements

This research is funded by an MRC/DFID African Research Leader Fellowship (MR/S005293/1: IMOA, CNM, RC, and AS). IMOA and JAGS have received grants from the Gavi, the Vaccine Alliance. JA GS is funded by a Wellcome Trust Senior Research Fellowship (214320) and the NIHR Health Protection Research Unit in Immunisation. DJN is funded by the Department of International Development and Wellcome [220985/z/20/z]. We thank Dr. Laura Hammitt, Ms. Angela Karani, the residents of the Kilifi Health and Demographic Surveillance System and the dedicated team of fieldworkers, administrative staff, clinicians, and laboratory staff who worked on this study. We acknowledge the continued collaboration of colleagues and the leadership in the Kilifi County Department of Health. This report is published with the permission of the Director of the Kenya Medical Research Institute

This research was partly funded by the Bill & Melinda Gates Foundation (INV-003174: MJ). BMGF (OPP1157270: KA). This project has received funding from the European Union’s Horizon 2020 research and innovation programme - project EpiPose (101003688: MJ). This research was partly funded by the National Institute for Health Research (NIHR) using UK aid from the UK Government to support global health research. The views expressed in this publication are those of the author(s) and not necessarily those of the NIHR or the UK Department of Health and Social Care (16/137/109: MJ; NIHR200929: MJ; NIHR Global Health Research Unit on Mucosal Pathogens: JO). Wellcome Trust (208812/Z/17/Z: SFlasche; 210758/Z/18/Z: SFunk).

The following authors were part of the Centre for Mathematical Modelling of Infectious Disease 2019-nCoV working group. Each contributed in processing, cleaning and interpretation of data, interpreted findings, contributed to the manuscript, and approved the work for publication: James D Munday, Carl A B Pearson, Simon R Procter, Oliver Brady, David Simons, Rachel Lowe, W John Edmunds, Katharine Sherratt, Rosanna C Barnard, Alicia Rosello, Adam J Kucharski, Fiona Yueqian Sun, Nikos I Bosse, Petra Klepac, Yang Liu, Kiesha Prem, Gwenan M Knight, Akira Endo, Sam Abbott, Emily S Nightingale, Thibaut Jombart, Jon C Emery, Georgia R Gore-Langton, Joel Hellewell, James W Rudge, Hamish P Gibbs, Kathleen O’Reilly, Kevin van Zandvoort, Yung-Wai Desmond Chan, Damien C Tully, Anna M Foss, Christopher I Jarvis, Katherine E. Atkins, Samuel Clifford, Matthew Quaife, Billy J Quilty, Rein M G J Houben, Rosalind M Eggo, Graham Medley, Sophie R Meakin, Timothy W Russell, Nicholas G. Davies, Charlie Diamond, Arminder K Deol, C Julian Villabona-Arenas, Stéphane Hué, Megan Auzenbergs, Quentin J Leclerc, Amy Gimma.

The following funding sources are acknowledged as providing funding for the working group authors. Alan Turing Institute (AE). BBSRC LIDP (BB/M009513/1: DS). This research was partly funded by the Bill & Melinda Gates Foundation (INV-001754: MQ; INV-003174: KP, YL; NTD Modelling Consortium OPP1184344: CABP, GFM; OPP1180644: SRP;OPP1183986: ESN; OPP1191821: KO’R, MA). DFID/Wellcome Trust (Epidemic Preparedness Coronavirus research programme 221303/Z/20/Z: CABP, KvZ). DTRA (HDTRA1-18-1-0051: JWR). Elrha R2HC/UK DFID/Wellcome Trust/This research was partly funded by the National Institute for Health Research (NIHR) using UK aid from the UK Government to support global health research. The views expressed in this publication are those of the author(s) and not necessarily those of the NIHR or the UK Department of Health and Social Care (KvZ). ERC Starting Grant (#757699: JCE, MQ, RMGJH). This project has received funding from the European Union’s Horizon 2020 research and innovation programme - project EpiPose (101003688: KP, PK, RCB, WJE, YL). This research was partly funded by the Global Challenges Research Fund (GCRF) project ‘RECAP’ managed through RCUK and ESRC (ES/P010873/1: AG, CIJ, TJ). HDR UK (MR/S003975/1: RME). Nakajima Foundation (AE). NIHR (16/136/46: BJQ; 16/137/109: BJQ, CD, FYS, YL; Health Protection Research Unit for Immunisation NIHR200929: NGD; Health Protection Research Unit for Modelling Methodology HPRU-2012-10096: TJ; PR-OD-1017-20002: AR, WJE). Royal Society (Dorothy Hodgkin Fellowship: RL; RP\EA\180004: PK). UK DHSC/UK Aid/NIHR (ITCRZ 03010: HPG). UK MRC (LID DTP MR/N013638/1: GRGL, QJL; MC_PC_19065: AG, NGD, RME, SC, TJ, WJE, YL; MR/P014658/1: GMK). Authors of this research receive funding from UK Public Health Rapid Support Team funded by the United Kingdom Department of Health and Social Care (TJ). Wellcome Trust (206250/Z/17/Z: AJK, TWR; 206471/Z/17/Z: OJB; 208812/Z/17/Z: SC; 210758/Z/18/Z: JDM, JH, KS, NIB, SA, SRM). No funding (AKD, AMF, CJVA, DCT, KEA, SH, YWDC).

---

**Figure S1.**
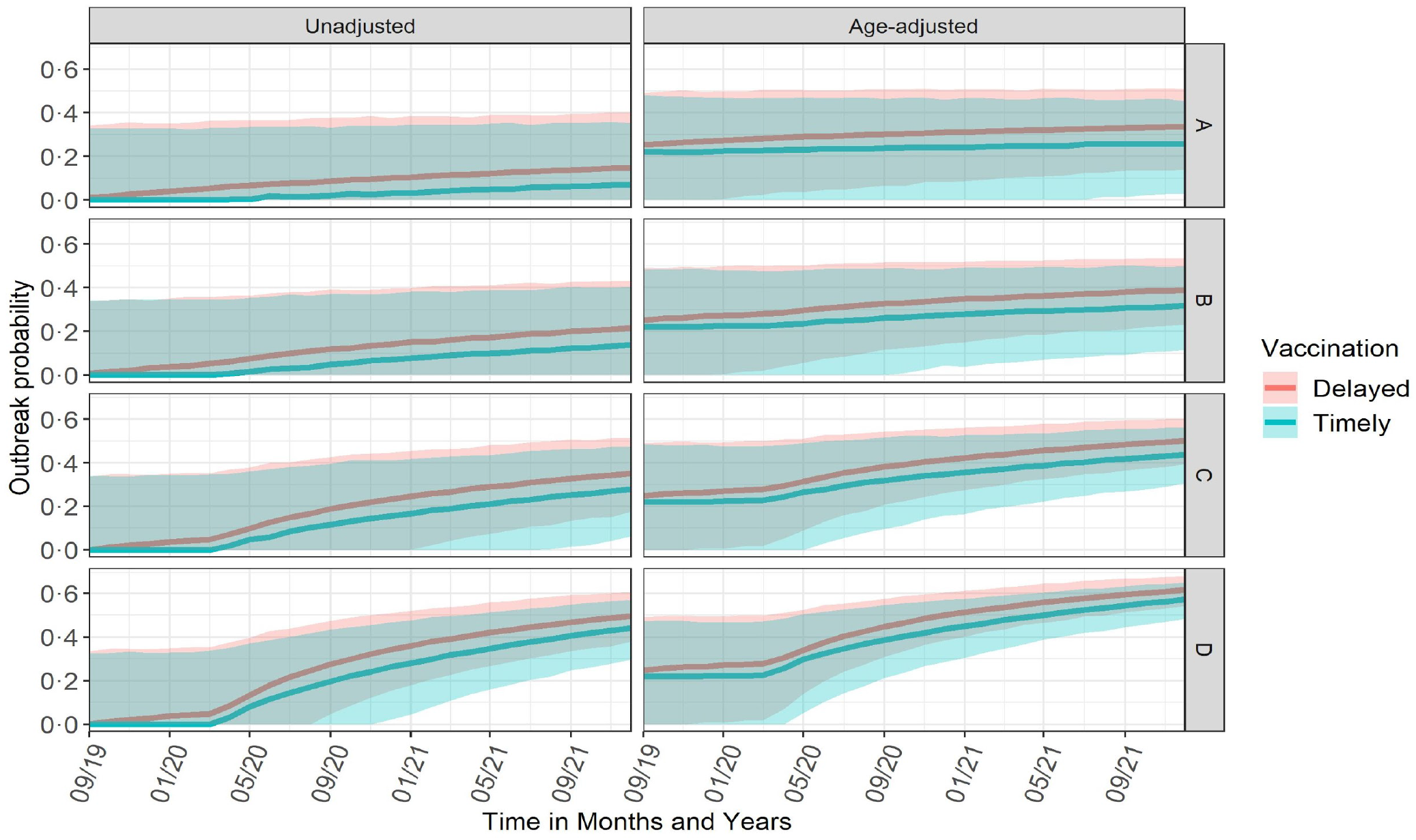
Probability of a large measles outbreak sparked by a single infected individual for timely and delayed vaccination.

**Figure S2.**
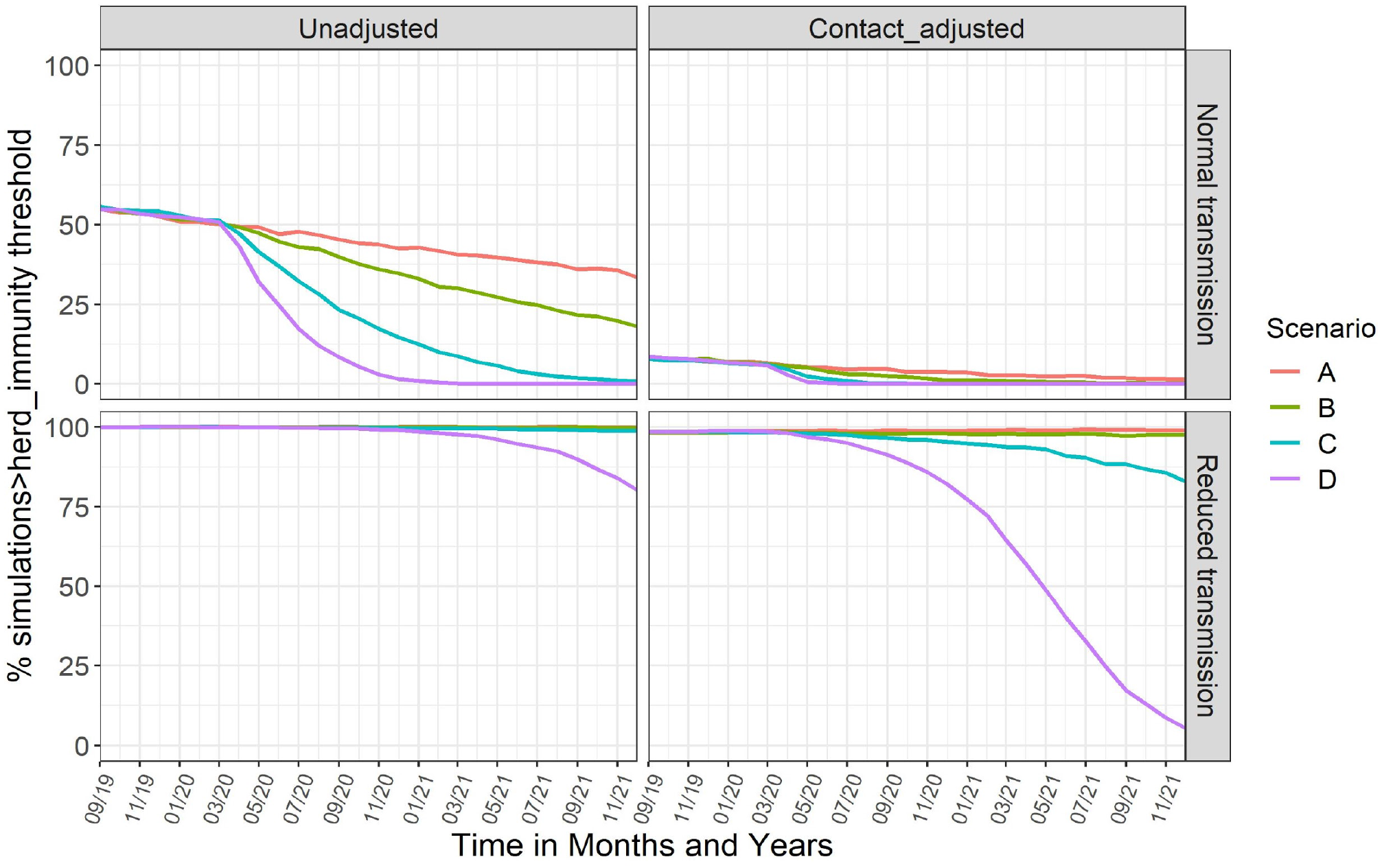
Percentage of simulations with proportion immune > herd immunity threshold.

**Figure S3.**
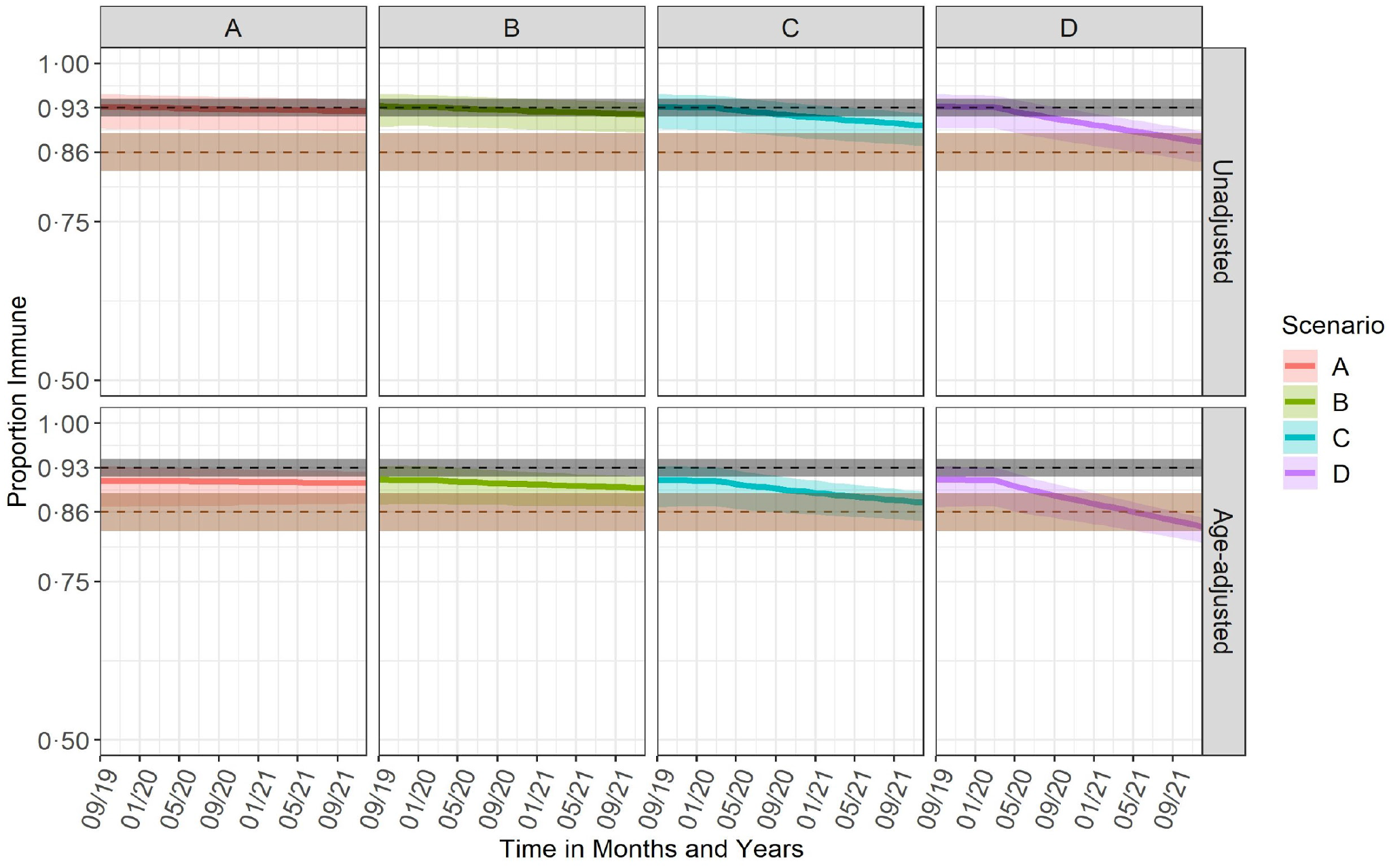
Monthly projected unadjusted and contact adjusted immunity profiles from September 2019 to December 2021. The changes in coverage took effect in April 2020. The black line shows the herd immunity threshold for measles before the COVID-19 pandemic 0·93 (0·92 to 0·94) and the brown line shows the herd immunity threshold during COVID-19 pandemic, 0·86 [0·83-0·89], assuming the lockdown measures are still in effect

**Figure S4.**
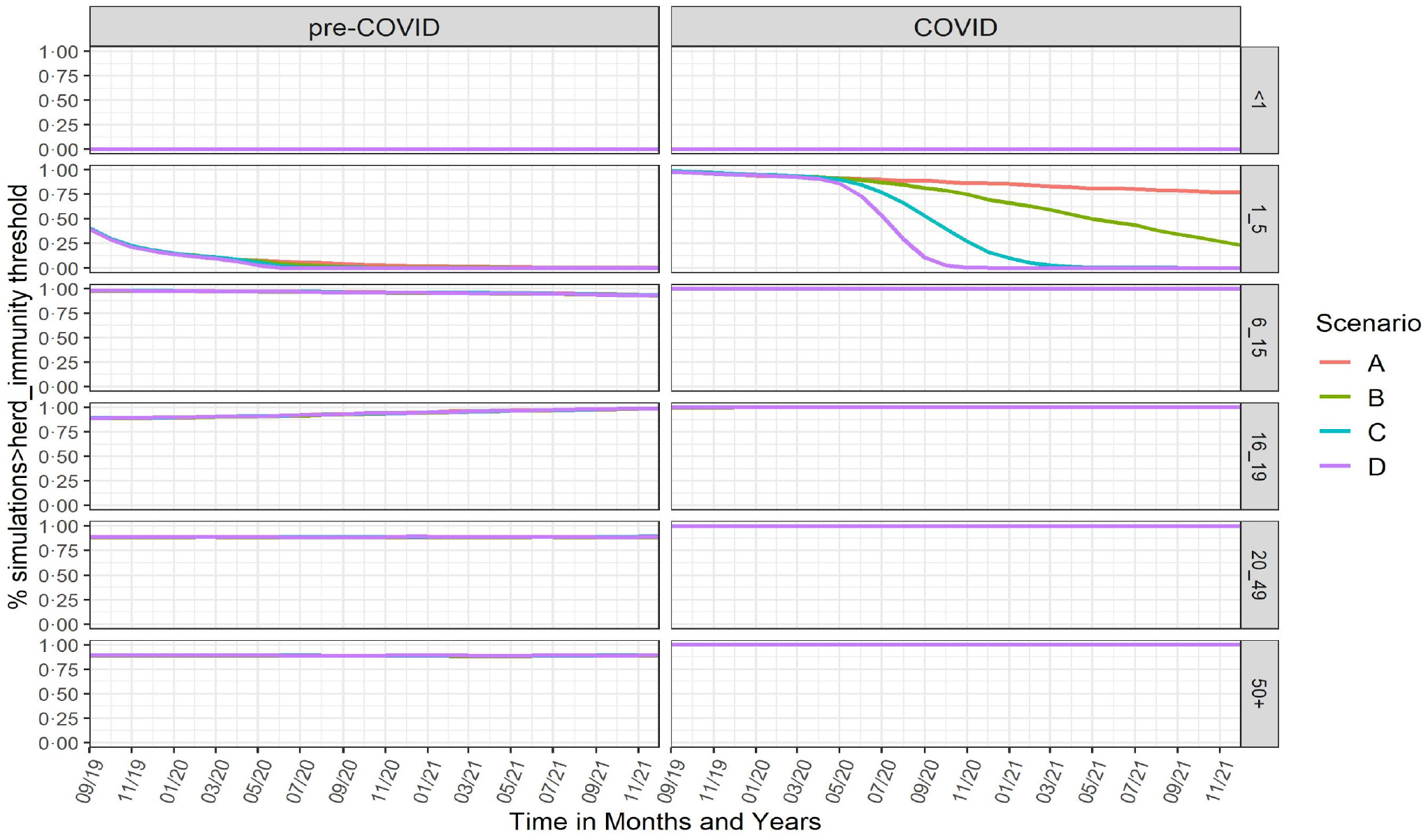
Percentage of simulations greater than herd immunity threshold.

